# Proximity-mapped whole-genome sequencing for characterization of repeat expansions and variants in highly homologous genomic regions

**DOI:** 10.64898/2026.10.11.26365260

**Authors:** Yuxiang Lin, Li Zhang, Yuzhen Qiang, Lu Yang, Xuefeng Li, Kaiyu Wang

**Affiliations:** Shanghai Fujun Medical Laboratory Co Ltd, Shanghai, China

**Keywords:** Proximity-mapped WGS, TruPath Genome, Short tandem repeats, Pseudogenes, Haplotype phasing, Clinical genomics

## Abstract

**Background:** Short-read whole-genome sequencing (sWGS) has limitations in characterizing tandem-repeat expansions and variants in highly homologous or pseudogene-associated regions. Long-read sequencing can address some of these challenges but often benefits from high-molecular-weight DNA. TruPath Genome incorporates spatial proximity information into a short-read sequencing framework. This study evaluated its performance in characterizing selected complex genomic variants using archived clinical DNA specimens.

**Methods:** Ten archived peripheral blood DNA specimens were analyzed, comprising seven cases with disease-associated short tandem repeat (STR) expansions and three cases involving homologous or pseudogene-associated loci (*STRC, SMN1/SMN2*, and *CYP21A2*). TruPath results were compared with previously available orthogonal assay results, including triplet repeat-primed PCR (TP-PCR), multiplex ligation-dependent probe amplification (MLPA), quantitative PCR (qPCR), and sWGS-based ExpansionHunter analysis.

**Results:** TruPath generated repeat-length estimates across the evaluated STR loci. For two *FMR1* full-mutation alleles, TruPath estimated 284 and 295 CGG repeats, respectively, while TP-PCR reported expansions exceeding 200 repeats. In a *DMPK* case, TruPath estimated an expanded allele of 1,203 CTG repeats, compared with a TP-PCR result exceeding 100 repeats. At the *STRC* and *SMN1/SMN2* loci, TruPath findings were consistent with available orthogonal results for deletion and copy-number status. At the *CYP21A2* locus, TruPath reconstructed the RCCX gene arrangement and assigned the I2G variant and a chimeric allele to different haplotypes.

**Conclusions:** In this small series of archived clinical DNA specimens, TruPath Genome enabled characterization of selected repeat expansions and homologous or pseudogene-associated variants. The findings support further evaluation of proximity-mapped sequencing as a complementary approach for complex genomic regions, particularly when conventional short-read sequencing is limited.

## 1. Introduction

Short-read whole-genome sequencing (sWGS) is increasingly used in the diagnosis of rare genetic disorders because it enables genome-wide assessment of multiple variant classes, including single-nucleotide variants (SNVs), small insertions and deletions (indels), copy-number variants (CNVs), and selected structural variants (SVs) [1–4]. However, its ability to characterize complex genomic variation is limited by short read lengths, particularly in repetitive regions and loci containing highly homologous paralogs or pseudogenes, where sequence ambiguity can compromise variant detection, copy-number assessment, and haplotype assignment [5, 6].

Tandem-repeat expansions are an important class of variation that remains challenging for sWGS. Pathogenic expansions in genes such as *FMR1, DMPK, FXN*, and *C9orf72* can extend to hundreds or thousands of repeat units and vary substantially in size [7–9]. Conventional molecular assays, including capillary electrophoresis and Southern blotting, remain important for clinical testing but have different limitations in sizing very large expansions, quantifying repeat lengths, and processing samples [10, 11]. Although bioinformatic tools such as ExpansionHunter enable repeat genotyping from sWGS data, their performance can be limited for large expansions that exceed the effective sizing range of short-read approaches [12–14]. The highly GC-rich *FMR1* CGG-repeat region is particularly challenging because of reduced sequencing representation, while large *DMPK* CTG expansions may be difficult to characterize quantitatively. These limitations can affect the molecular characterization and interpretation of repeat-associated disorders.

A related challenge arises at loci containing highly homologous genes and pseudogenes, including *STRC/STRCP1, SMN1/SMN2*, and *CYP21A2/CYP21A1P* [15–17]. Sequence similarity and complex gene arrangements can complicate the assignment of reads and variants to the correct gene copy, potentially leading to missed variants, ambiguous variant localization, or inaccurate copy-number estimates [18–20]. In these regions, establishing the genomic origin and phase of a variant may be as important as detecting the variant itself. Clinical characterization therefore often requires complementary methods, such as multiplex ligation-dependent probe amplification (MLPA), quantitative PCR (qPCR), long-range PCR with Sanger sequencing, or targeted long-read sequencing [15, 16, 21–23]. The need for complementary testing highlights the difficulty of resolving sequence variation within highly homologous genomic regions.

TruPath Genome (Illumina) incorporates spatial proximity information into a short-read sequencing workflow, providing genomic context beyond that available from individual short reads. During library preparation, DNA is processed using transposase-based fragmentation, while proximity-associated information is retained for downstream analysis. Previous evaluations have examined its application to genome-wide variant detection, long-range phasing, and structural variant characterization, with reported performance for SNVs and small indels comparable to conventional PCR-free WGS [24]. However, its ability to characterize large tandem-repeat expansions and resolve variants at highly homologous or pseudogene-associated loci in clinical specimens remains insufficiently characterized.

In this study, we evaluated TruPath Genome using 10 archived peripheral blood DNA specimens with previously characterized complex genomic variants. Seven specimens harbored disease-associated STR expansions across 13 loci, and three involved the homologous or pseudogene-associated regions *STRC/STRCP1, SMN1/SMN2*, and *CYP21A2/CYP21A1P*. TruPath results were compared with previously available orthogonal molecular assay results and conventional sWGS-based ExpansionHunter estimates, where applicable. We assessed repeat-length estimation, locus-specific copy-number characterization, and the resolution of complex gene arrangements and variant phase, with particular attention to performance in archived DNA specimens without a dedicated ultra-high-molecular-weight DNA extraction workflow.

## 2. Materials and methods

### 2.1 Study Design and Sample Selection

This study included 10 archived peripheral blood DNA specimens originally submitted to Shanghai FuJun Genetics Technology Co., Ltd. for genetic testing. All specimens were de-identified before analysis. Written informed consent was obtained from participants or their legal guardians for the use of residual specimens in method validation and subsequent research. The specimens had been stored at −20°C for up to two years, and some had undergone multiple freeze–thaw cycles. The specimens were classified into two groups according to the type of genomic variation and the associated analytical challenges: (1) tandem-repeat expansions and (2) variants involving homologous or pseudogene-associated loci.

The repeat-expansion group comprised seven specimens (TP01–TP07) covering 13 disease-associated STR loci in 13 genes: *AR, ATN1, C9orf72, DMPK, FMR1, FXN, HTT, ATXN1, ATXN2, ATXN3, ATXN7, ATXN10*, and *ATXN8OS*. TruPath repeat-length estimates were compared with previously available TP-PCR results or conventional short-read WGS-based ExpansionHunter estimates, as applicable. Three specimens represented particularly challenging expansion cases: two *FMR1* full-mutation alleles (>200 CGG repeats) and one *DMPK* expansion (>100 CTG repeats), for which TP-PCR confirmed expansion beyond the reported threshold but did not provide exact repeat lengths. The remaining loci were evaluated by comparison with available quantitative reference results.

The second group comprised three specimens (TP08–TP10) involving homologous or pseudogene-associated loci: a deletion involving *STRC*, a deletion resulting in a single remaining *SMN1* copy with two copies of *SMN2*, and complex variation in the *CYP21A2/CYP21A1P* RCCX region involving the intronic variant c.293-13A/C>G (I2G) and a chimeric *CYP21A1P/CYP21A2* allele (CH-1). Previously available orthogonal results, including qPCR and MLPA, were used for comparison where applicable. The study therefore focused on the characterization of selected known variants rather than a comprehensive assessment of diagnostic sensitivity or specificity.

### 2.2 DNA quantification and integrity assessment

Genomic DNA was extracted from EDTA-anticoagulated whole blood using the MagMAX™ DNA Multi-Sample Ultra 2.0 Kit (Thermo Fisher Scientific, USA) following standard magnetic-bead protocols without dedicated high-molecular-weight (HMW) extraction workflows. DNA concentration was measured using a Qubit 4.0 fluorometer (Thermo Fisher Scientific, USA). DNA integrity was assessed using an Agilent TapeStation 2200 system, providing the DNA Integrity Number (DIN) and the proportions of DNA fragments >10 kb and >60 kb based on electrophoretic signal analysis. Sample quality metrics were recorded to evaluate their impact on TruPath proximity-linked parameters. Specimens were not excluded based solely on DNA integrity measurements.

### 2.3 TruPath Genome library preparation and sequencing

TruPath Genome library preparation and sequencing were performed using the automated TruPath workflow (Illumina) according to the manufacturer’s instructions. Briefly, 350 ng of genomic DNA was loaded per sample, after which library preparation and sequencing proceeded through the automated workflow. The library preparation process incorporated on-bead, transposase-based fragmentation without prior mechanical shearing or gel-based size selection. Sequencing was performed on a NovaSeq X Plus platform (Illumina) using 150-bp paired-end reads with the TruPath proximity module enabled. The target mean autosomal sequencing depth was ≥50×.

### 2.4 Data processing and quality metrics

Secondary analysis was performed using the DRAGEN server (v4.5.4, Illumina) with proximity mode enabled. DRAGEN fitted a spatial proximity linking model on reads originating from the same DNA molecule on the flow cell, computing Phred-scaled linkage probabilities and tagging reconstructed template molecules. For homologous and pseudogene regions, the Multi-Region Joint Detection (MRJD) algorithm was applied to enable copy- and locus-specific variant assignment. For STR loci, proximity-guided placement of in-repeat reads (IRRs) was performed across the 13 targeted repeat genes. Quality control metrics included standard WGS metrics (mean coverage, % >20× coverage, % Q30 bases, total input reads, mapping rate, unique mapping rate, proper pairing rate, and median insert size) and TruPath-specific proximity metrics (Fit RMSE, Q25 proximity rate, Q25 proximity coverage, P75 template size, and phase block NG50).

### 2.5 TP-PCR, MLPA, and qPCR

Triplet repeat-primed PCR (TP-PCR) was performed to detect trinucleotide repeat expansions in FMR1, DMPK, ATXN2, and ATXN3. Flanking primers combined with repeat-anchored primers were amplified, and capillary electrophoresis was used to identify expanded alleles and estimate repeat counts. Orthogonal validation of variants in SMN1 and CYP21A2 was conducted using multiplex ligation-dependent probe amplification (MLPA; MRC Holland, Amsterdam, Netherlands) with P060 and P050 probe mixes, respectively. Variants in STRC were validated using quantitative PCR (qPCR). Due to pseudogene sequence homology (STRCP1), STRC-specific qPCR primers were restricted to exons 16 and 24 (E16F: ACCTTACCCTCAGGAATCTG, E16R: TATCCCCCATTCCATATCAC; E24F: GCAAGACTTAAAACCCAGCAGTA, E24R: CTAGACCTATTAAGAGCCTACCA).

## 3. Results

### 3.1 DNA Quality and Quality Control Parameters

DNA quality and sequencing performance were assessed for all 10 specimens (Table 1 and Supplementary Table 1). The archived specimens were processed using the Thermo Fisher MagMAX DNA Multi-Sample Ultra kit without a dedicated ultra-high-molecular-weight DNA extraction workflow. DNA integrity number (DIN) ranged from 7.0 to 9.0. The proportion of DNA fragments >10 kb ranged from 60.64% to 90.58%, and that of fragments >60 kb ranged from 13.49% to 30.56%. Owing to long-term storage, the specimens showed considerable variation in DNA integrity and the proportion of long DNA fragments. Several specimens were close to the lower QC boundary, with >10% of DNA fragments >60 kb used as the minimum criterion reported for proximity-mapped sequencing studies [24].

**Table 1.** DNA quality and TruPath Genome sequencing metrics for the 10 study samples.

| **Sample ID** | **DIN** | **>10kb (%)** | **>60kb (%)** | **Fit RMSE** | **Q25 Proximity Rate** | **Q25 Proximity Coverage** | **P75 Template Size ( bp )** | **Phase Block NG50 ( bp )** |
| --- | --- | --- | --- | --- | --- | --- | --- | --- |
| TP01 | 8.3 | 82.71 | 18.24 | 1.72 | 18.42% | 16.33 | 6,254 | 122,880 |
| TP02 | 8.5 | 82.71 | 30.56 | 0.64 | 30.15% | 20.07 | 9,071 | 176,128 |
| TP03 | 7 | 61.32 | 15.29 | 0.55 | 21.48% | 18.09 | 10,426 | 229,376 |
| TP04 | 8.0 | 81.8 | 25.25 | 1.16 | 24.48% | 20.06 | 8,328 | 151,552 |
| TP05 | 7.1 | 60.64 | 13.49 | 1 | 18.95% | 16.23 | 6,841 | 118,784 |
| TP06 | 8.1 | 78.16 | 17.96 | 2.07 | 16.01% | 13.51 | 5,214 | 86,016 |
| TP07 | 8.1 | 77.65 | 19.34 | 0.33 | 21.89% | 19.16 | 7,647 | 139,264 |
| TP08 | 7.4 | 66.26 | 28.87 | 0.4 | 31.73% | 26.34 | 16,096 | 299,008 |
| TP09 | 7.8 | 77.93 | 20.23 | 0.92 | 21.16% | 17.31 | 8,111 | 139,264 |
| TP10 | 9 | 90.58 | 30.04 | 0.45 | 35.10% | 26.09 | 14,072 | 217,088 |

Summary of DNA integrity parameters and TruPath proximity-mapped sequencing metrics for the archived samples. DIN, DNA integrity number; Fit RMSE, root-mean-square error of proximity model fit.

Conventional WGS metrics were relatively consistent across specimens. Mean autosomal coverage ranged from 49.33× to 67.78×, with 93.17–94.21% of bases covered at ≥20×. Q30 values ranged from 90.63% to 92.19%. The number of input reads ranged from 1.58 to 1.88 billion, while mapped-read, unique-read, and proper-pair rates were 95.67–96.91%, 84.36–92.91%, and 91.44–93.20%, respectively. Median insert sizes ranged from 188 to 217 bp. Overall, conventional WGS metrics showed limited inter-sample variation, although TP02 had relatively lower mean autosomal coverage and input read count.

In contrast, TruPath-specific proximity metrics showed greater inter-sample variation. Q25 proximity rate ranged from 16.01% to 35.10%, meeting the MJDR reference threshold of >16% in all specimens, while Q25 proximity coverage ranged from 13.51× to 26.34×. Fit RMSE ranged from 0.33 to 2.07. P75 reconstructed template size ranged from 5,214 to 16,096 bp, and phase-block NG50 ranged from 86,016 to 299,008 bp. The relatively low and variable proximity metrics were consistent with differences in DNA integrity and the availability of long DNA fragments. Specimens with higher proportions of DNA fragments >60 kb generally showed larger reconstructed template sizes and longer phase blocks. For example, TP08 and TP10, with >60-kb fractions of 28.87% and 30.04%, respectively, showed the largest P75 template sizes (16,096 and 14,072 bp) and the highest phase-block NG50 values (299,008 and 217,088 bp).

### 3.2 Detection of STR Expansions and Repeat-Length Estimation

To evaluate the ability of TruPath to detect disease-associated STR expansions and estimate repeat lengths, 13 STR loci were assessed across seven specimens (TP01–TP07; Table 2). At several loci, conventional short-read WGS (sWGS) showed limited coverage or mapping performance, particularly in GC-rich or repetitive regions such as *FMR1*, limiting reliable characterization of expanded alleles. TruPath provided proximity-linked information that supported detection and repeat-length estimation at these loci.

**Table 2.**
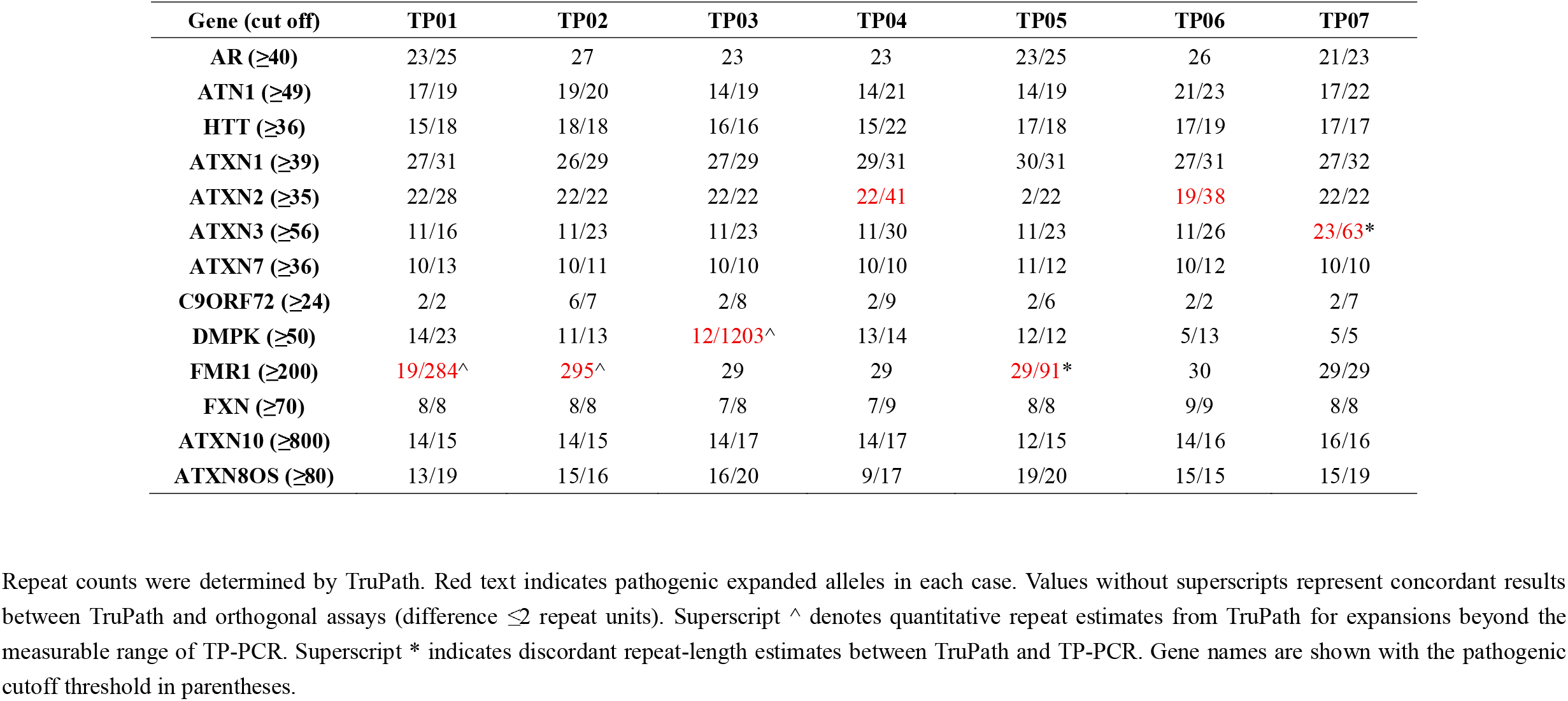
STR repeat counts across 13 disease-associated tandem repeat loci in seven samples.

For large expansions, TP-PCR identified full-mutation *FMR1* alleles (>200 CGG repeats) in TP01 and TP02, whereas TruPath estimated 284 and 295 CGG repeats, respectively. Because TP-PCR reported these alleles only as >200 repeats, quantitative agreement between the two methods could not be directly assessed. At *DMPK* in TP03, TP-PCR detected an expansion of >100 CTG repeats, while TruPath estimated one expanded allele of 1,203 CTG repeats and one normal-sized allele of 12 repeats, providing a quantitative estimate beyond the reporting range of the reference assay (Supplementary Fig. 1).

For expanded alleles within the quantitative range of the reference assays, some differences in repeat-length estimates were observed. In TP05, TruPath estimated *FMR1* alleles of 29/91 repeats, compared with 29/78 repeats by TP-PCR. At *ATXN3* in TP07, TruPath estimated 26/63 repeats versus 26/71 repeats by TP-PCR (Supplementary Fig. 1). In both specimens, the normal-sized alleles showed close agreement, whereas the differences were primarily observed for the expanded alleles. At *ATXN2*, TruPath estimates of 22/41 repeats in TP04 and 19/38 repeats in TP06 were concordant with the corresponding orthogonal results. Across the remaining evaluated loci, TruPath estimates were generally comparable with ExpansionHunter results from conventional sWGS, with differences of no more than two repeat units for most alleles.

Overall, TruPath detected and estimated repeat lengths across a range of STR loci and expansion sizes, including expansions for which the reference assay provided only a categorical or upper-bound result. For large expansions beyond the quantitative range of the reference assay, TruPath-derived repeat lengths should be regarded as estimates rather than independently validated repeat counts. Orthogonal testing remains appropriate when accurate repeat sizing is required for clinical interpretation.

### 3.3 Homologous and Pseudogene-Associated Loci: STRC, SMN1/SMN2, and CYP21A2

Homologous and pseudogene-associated regions can be challenging for conventional short-read WGS because of sequence similarity and ambiguous read mapping. Manual inspection of BAM files suggested improved read-level mapping at several pseudogene-associated loci, although the extent of improvement varied across regions (Supplementary Fig. 2). To further assess performance at clinically relevant loci, three regions were evaluated using TruPath MRJD analysis.

In TP08, TruPath identified a deletion involving the functional STRC locus while distinguishing it from the highly homologous pseudogene STRCP1 (Fig. 1A). Previously obtained qPCR results for selected exons, including exons 16 and 24, were consistent with the STRC deletion identified by TruPath (Fig. 1C). The qPCR assay targeted selected exons because of the high sequence homology in this region.

**Figure 1.**
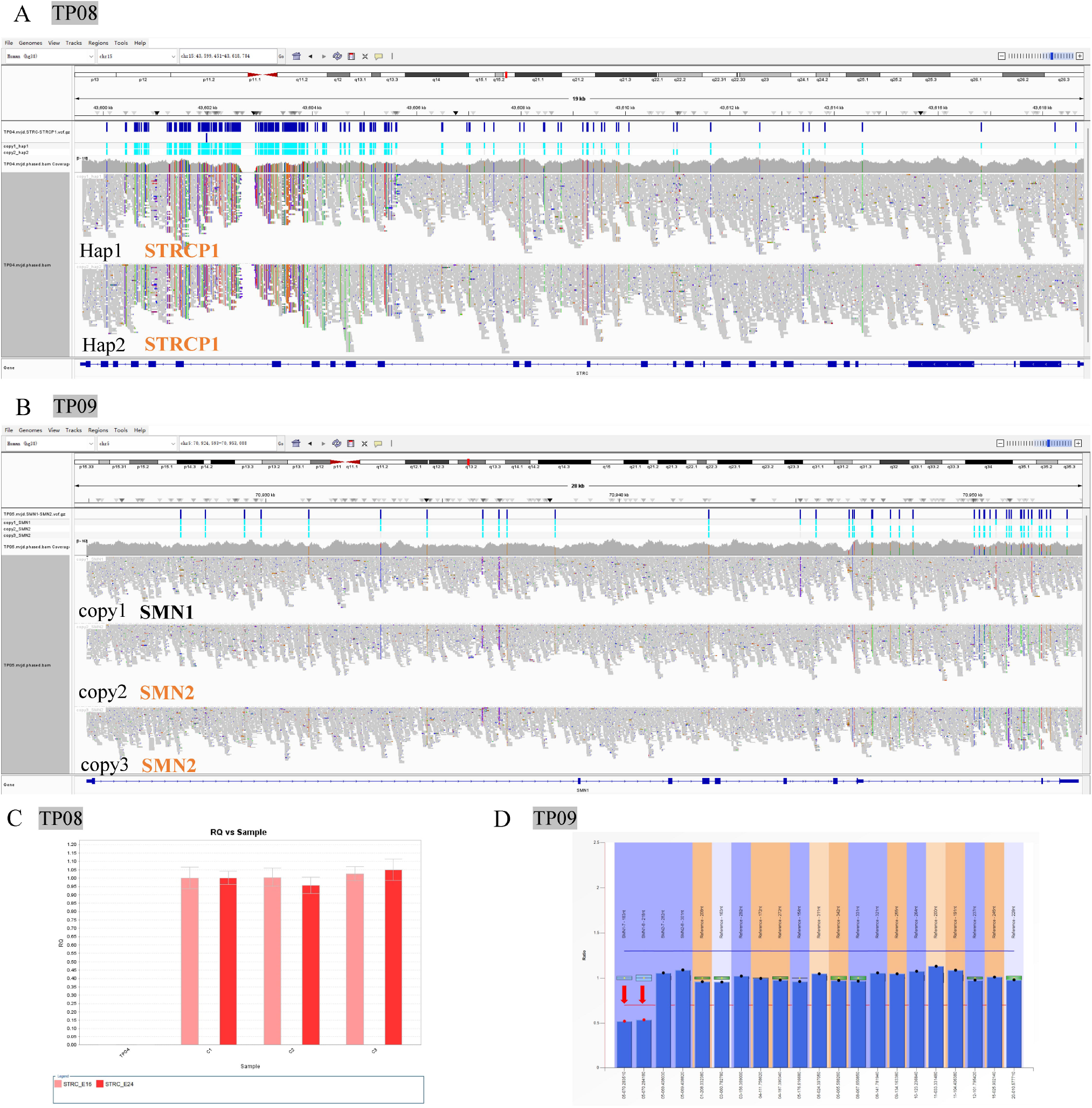
TruPath MRJD resolves copy number variants in regions with highly homologous paralogs. (A) IGV visualization of sample TP08 showing a deletion in the functional STRC gene, clearly discriminated from its homologous pseudogene STRCP1. Two haplotypes (Hap1 and Hap2) are displayed. (B) IGV view for sample TP09 showing copy-number reconstruction of SMN1 and SMN2. One copy of SMN1 and two copies of SMN2 were identified. (C) Quantitative PCR (qPCR) validation of the STRC deletion in TP08, targeting exons 16 and 24 of STRC. (D) MLPA validation of the SMN1/SMN2 copy number profile for TP09.

In TP09, TruPath determined the copy-number status of SMN1 and SMN2, identifying a deletion resulting in a single remaining SMN1 copy and two copies of SMN2 (Fig. 1B). These copy-number findings were concordant with MLPA results (Fig. 1D).

In TP10, TruPath MRJD analysis reconstructed four gene copies within the RCCX region and assigned them to two haplotypes. Haplotype 1 contained a *CYP21A2* copy carrying the intronic splice-site variant c.293-13A/C>G and a *CYP21A1P* copy. Haplotype 2 contained a *CYP21A1P* copy and a chimeric CYP21A1P/CYP21A2 allele designated CH-1 (Fig. 2A, C). Copy-number findings were concordant with MLPA (Fig. 2B). These results illustrate the potential of proximity-linked sequencing to characterize complex gene arrangements and variant phase at the RCCX locus.

**Figure 2.**
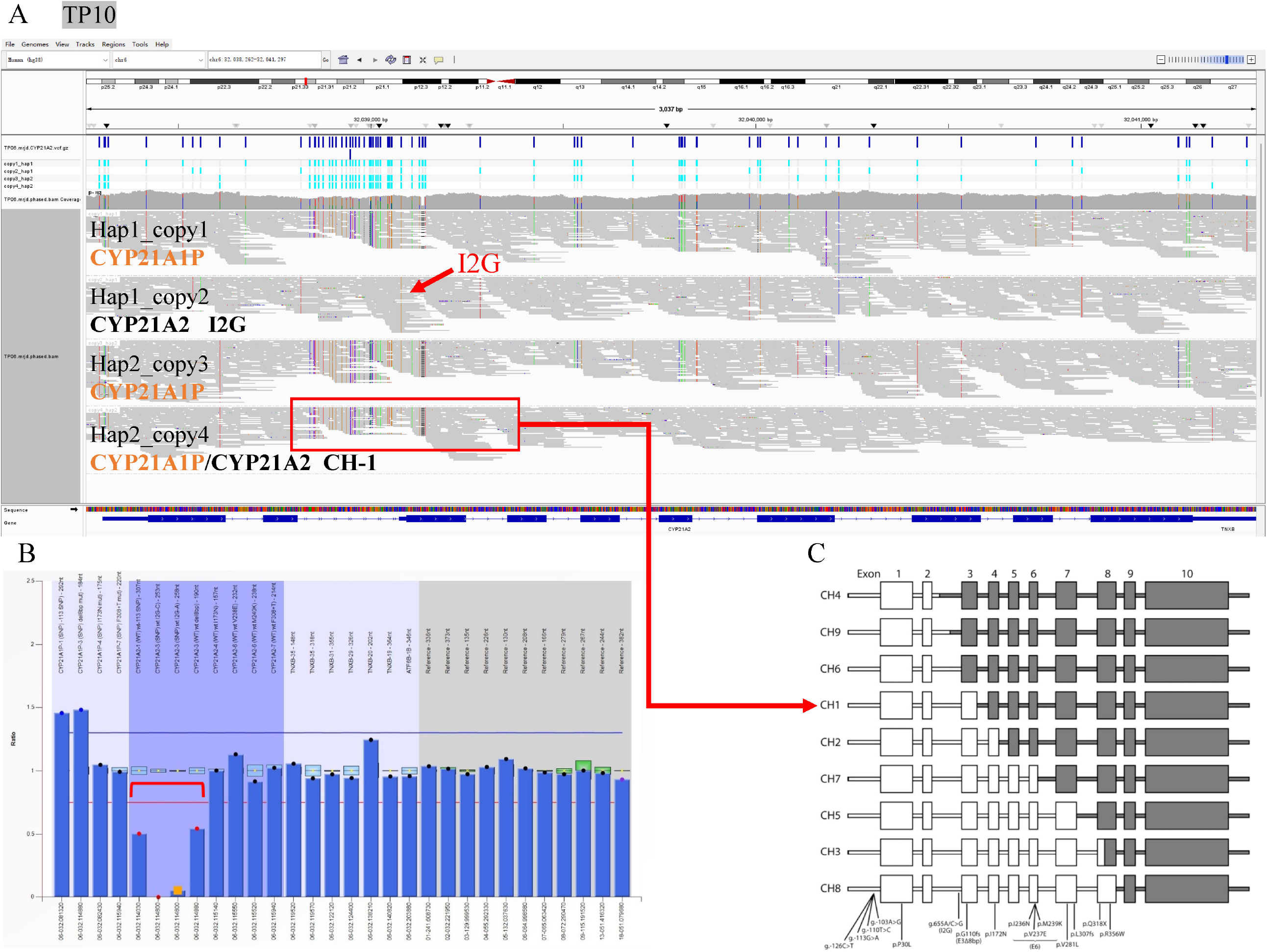
TruPath MRJD resolves complex structural variants and phasing in the RCCX region. (A) IGV visualization of sample TP10 showing four reconstructed copies within the RCCX region assigned to two haplotypes. Haplotype 1 includes one CYP21A1P copy and one CYP21A2 copy harboring the c.293-13A/C>G (I2G) splice-site variant. Haplotype 2 comprises one CYP21A1P copy and the chimeric CYP21A1P/CYP21A2 CH-1 fusion allele. The red arrow marks the I2G variant; the red box highlights the CH-1 chimeric fusion allele. (B) MLPA copy-number profile of TP10, confirming the copy number results identified by TruPath MRJD. (C) Schematic diagram illustrating the haplotype structure of the CH-1 chimeric allele in the RCCX region.

Overall, TruPath characterized copy-number changes at STRC and SMN1/SMN2 and resolved the gene arrangement and variant phase at the RCCX locus. The agreement with previously available orthogonal results provides independent support for the corresponding findings, while the BAM-file observations suggest potential improvements in read mapping at selected pseudogene-associated regions.

## 4. Discussion

In this study, we evaluated proximity-mapped WGS for characterizing pathogenic repeat expansions and variants in highly homologous or pseudogene-associated loci. Across the evaluated specimens, TruPath provided repeat-length estimates at 13 disease-associated STR loci and characterized selected variants involving *STRC, SMN1/SMN2*, and *CYP21A2*. These findings suggest that proximity information can provide additional genomic context in regions where conventional short-read sequencing is challenged by repetitive sequences or high sequence homology.

The repeat-expansion analysis illustrates the potential value of this approach for selected loci that are difficult to characterize by conventional sWGS. For two *FMR1* full-mutation alleles, TruPath estimated 284 and 295 CGG repeats, whereas TP-PCR reported expansions exceeding 200 repeats. Similarly, at *DMPK*, TruPath estimated an expanded allele of 1,203 CTG repeats, compared with a TP-PCR result exceeding 100 repeats. These results illustrate the potential of proximity-mapped sequencing to provide quantitative estimates beyond the reporting thresholds of the corresponding TP-PCR assays. However, because the reference assays did not provide exact repeat counts for these large expansions, the estimates could not be independently verified. Differences between TruPath and TP-PCR estimates were also observed for some expanded alleles, including those in *FMR1* and *ATXN3*. Accordingly, repeat-length estimates, particularly for large expansions, should be interpreted cautiously, and orthogonal testing remains important when precise sizing is required for clinical interpretation.

At homologous and pseudogene-associated loci, proximity information provided complementary information on gene-specific copy number and genomic structure. At the *STRC/STRCP1* locus, TruPath characterized a deletion involving the functional *STRC* gene, consistent with previously available qPCR results for selected exons. At the *SMN1/SMN2* locus, TruPath identified a copy-number state consistent with previously available MLPA results. At the RCCX locus in TP10, MRJD reconstructed a four-copy gene arrangement and assigned the *CYP21A2* intronic variant c.293-13A/C>G (I2G) and the CH-1 chimeric allele to different haplotypes. These findings illustrate the potential value of proximity-based analysis at loci where interpretation depends not only on variant detection or copy-number assessment but also on distinguishing highly homologous gene copies and resolving haplotype structure. Nevertheless, only one specimen was evaluated for each of these loci, and the findings do not establish performance across the full range of alleles encountered in clinical practice.

An additional practical consideration is that the archived DNA specimens were processed without a dedicated HMW DNA extraction workflow. DNA integrity and TruPath proximity metrics varied among specimens, while the evaluated findings were generally consistent with the corresponding previously available reference results. Descriptively, specimens with larger proportions of long DNA fragments tended to show higher proximity-related metrics. However, the study was not designed to quantify the relationship between DNA integrity and analytical performance, and the small sample size precludes defining a DNA quality threshold for successful clinical testing. Further evaluation using specimens with systematically characterized DNA integrity will be needed to determine how DNA quality affects proximity-based analysis.

From a diagnostic perspective, proximity-mapped WGS may offer a complementary means of investigating multiple classes of genomic variation within a genome-wide assay [24]. Repeat expansions, pseudogene-associated variants, copy-number changes, and phasing challenges often require different targeted methods in clinical genetics [19, 20, 25, 26]. Proximity-based analysis may provide additional information for selected difficult loci within a broader WGS workflow, potentially reducing the need for separate targeted assays in some cases. However, these potential workflow advantages were not directly evaluated in the present study. Established targeted methods remain important when definitive repeat sizing, locus-specific confirmation, or other clinically validated measurements are required.

Several limitations should be acknowledged. First, this was a small feasibility study involving 10 archived specimens with previously characterized variants. The number of specimens per locus was insufficient to estimate sensitivity, specificity, or diagnostic accuracy. Second, the largest repeat expansions exceeded the quantitative reporting range of the corresponding TP-PCR assays, limiting independent verification of the TruPath-derived repeat-length estimates. Third, the study focused on selected known-positive specimens and did not assess negative specimens, mosaic expansions, intermediate or borderline repeat alleles, or the broader spectrum of structural variants encountered in clinical practice. Finally, although DNA integrity and proximity metrics varied among specimens, no systematic evaluation across a defined range of DNA integrity was performed. Larger studies incorporating positive and negative clinical specimens, independent reference methods capable of resolving repeat lengths and complex gene structures, and a broader range of variant types will be required to establish the analytical performance and clinical utility of this approach.

In conclusion, proximity-mapped WGS provided additional genomic context for characterizing selected repeat expansions and variants in highly homologous or pseudogene-associated loci. In this small feasibility study, TruPath generated repeat-length estimates beyond the reporting thresholds of the corresponding TP-PCR assays and provided gene- and haplotype-level information at selected complex genomic regions. These findings support further evaluation of proximity-mapped WGS as a complementary approach for investigating inherited disorders involving genomic regions that are difficult to resolve using conventional short-read sequencing.

## Data Availability

All data produced in the present work are contained in the manuscript

## Patient consent statement

Written informed consent was obtained from participants or their legal guardians for the use of residual specimens in method validation and subsequent research.

## Data availability statement

Data available on request due to privacy/ethical restrictions.

## Acknowledgements

We gratefully acknowledge Lu Zhang of Illumina GC for her valuable guidance and technical support with the data analysis and interpretation in this study.

## Author contributions

Yuxiang Lin: Conceptualization, Methodology, Formal analysis, Investigation, Writing-original draft.

Li Zhang: Investigation, Data curation.

Yuzhen Qiang: Investigation, Sample processing, Validation.

Lu Yang: Investigation, Data curation, Validation.

Xuefeng Li: Resources, Data curation.

Kaiyu Wang^*^: Conceptualization, Resources, Supervision, Writing‑review & editing, Project administration.

---

Supplementary Figure 1. TP-PCR capillary electrophoresis profiles of repeat expansion samples. TP-PCR capillary electropherograms of samples TP01, TP02, TP03, TP05 and TP07 for FMR1, DMPK, and ATXN3 repeat expansions.

Supplementary Figure 2. IGV comparison between conventional short-read WGS and TruPath Genome sequencing in homologous and pseudogene-containing genomic regions. (A,B) IGV snapshots of conventional short-read WGS for HBA1/HBA2 and CYP21A2. (C,D) IGV snapshots of TruPath Genome for HBA1/HBA2 and CYP21A2.

## References

[1] Turro E, Astle WJ, Megy K, et al. Whole-genome sequencing of patients with rare diseases in a national health system. 2020; 583:96–102.

[2] Smedley D, Smith KR, Martin A, et al. 100,000 Genomes Pilot on Rare-Disease Diagnosis in Health Care - Preliminary Report. 2021; 385:1868–1880.

[3] Lionel AC, Costain G, Monfared N, et al. Improved diagnostic yield compared with targeted gene sequencing panels suggests a role for whole-genome sequencing as a first-tier genetic test. 2018; 20:435–443.

[4] Goodwin S, McPherson JD, McCombie WRJNRG. Coming of age: ten years of next-generation sequencing technologies. 2016; 17:333–351.

[5] Chaisson MJP, Wilson RK, Eichler EEJNRG. Genetic variation and the de novo assembly of human genomes. 2015; 16:627–640.

[6] Ebert P, Audano PA, Zhu Q, et al. Haplotype-resolved diverse human genomes and integrated analysis of structural variation. 2021; 372:eabf7117.

[7] Hannan Ajjnrg. Tandem repeats mediating genetic plasticity in health and disease. 2018; 19:286–298.

[8] Monaghan KG, Lyon E, Spector Ebjgm. ACMG Standards and Guidelines for fragile X testing: a revision to the disease-specific supplements to the Standards and Guidelines for Clinical Genetics Laboratories of the American College of Medical Genetics and Genomics. 2013; 15:575–586.

[9] Udd B, Krahe R. The myotonic dystrophies: molecular, clinical, and therapeutic challenges. The Lancet Neurology 2012; 11:891–905.

[10] Seifert BA, Reddi HV, Kang BE, Bean LJH, Shealy A, Rose NCJGM. Myotonic dystrophy type 1 testing, 2024 revision: A technical standard of the American College of Medical Genetics and Genomics (ACMG). 2024; 26:101145.

[11] Kamsteeg E-J, Kress W, Catalli C, et al. Best practice guidelines and recommendations on the molecular diagnosis of myotonic dystrophy types 1 and 2. 2012; 20:1203–1208.

[12] Dolzhenko E, Vugt JJFA, Shaw RJ, et al. Detection of long repeat expansions from PCR-free wholegenome sequence data. 2017; 27:1895–1903.

[13] Tankard RM, Bennett MF, Degorski P, Delatycki MB, Lockhart PJ, Bahlo MJAJHG. Detecting Expansions of Tandem Repeats in Cohorts Sequenced with Short-Read Sequencing Data. 2018; 103:858–873.

[14] Ibañez K, Jadhav B, Zanovello M, et al. Increased frequency of repeat expansion mutations across different populations. 2024; 30:3357–3368.

[15] Chen X, Harting J, Farrow E, et al. Comprehensive SMN1 and SMN2 profiling for spinal muscular atrophy analysis using long-read PacBio HiFi sequencing. 2023; 110:240–250.

[16] Xiang J, Peng J, Sun X, et al. The Next Generation of Population-Based DFNB16 Carrier Screening and Diagnosis: STRC Copy-Number Variant Analysis from Genome Sequencing Data. 2023; 69:763–770.

[17] Carvalho B, Marques CJ, Santos-Silva R, Fontoura M, Carvalho D, Carvalho F. Congenital Adrenal Hyperplasia Due to 21-Hydroxylase Deficiency: An Update on Genetic Analysis of CYP21A2 Gene. Experimental and clinical endocrinology & diabetes : official journal, German Society of Endocrinology [and] German Diabetes Association 2021; 129:477–481.

[18] Costa-Roger M, Blasco-Pérez L, Gerin L, et al. Complex SMN Hybrids Detected in a Cohort of 31 Patients With Spinal Muscular Atrophy. 2024; 10:e200175.

[19] Moteki H, Nishio S-Y, Usami S-IJSR. Targeted long-read nanopore sequencing as a complementary approach for detecting STRC variants and distinguishing the STRCP1 pseudogene. 2025; 15:44261.

[20] Miranda MC, Oliveira ACM, Soares GJ, et al. Evolution of molecular diagnostic strategies for 21-hydroxylase deficiency: from classical methods to advanced genomic techniques. 2026; 17:1770274.

[21] Kleinle S, Scholz V, Benet-Pagés A, et al. Closing the Gap - Detection of 5q-Spinal Muscular Atrophy by Short-Read Next-Generation Sequencing and Unexpected Results in a Diagnostic Patient Cohort. 2023; 10:835–846.

[22] Li H, Zhu X, Yang Y, et al. Long-read sequencing: An effective method for genetic analysis of CYP21A2 variation in congenital adrenal hyperplasia. 2023; 547:117419.

[23] Liu Y, Chen M, Liu J, et al. Comprehensive Analysis of Congenital Adrenal Hyperplasia Using Long-Read Sequencing. Clin Chem 2022; 68:927–939.

[24] Bertoli L, Orlandi M, Timis AI, et al. Illumina TruPath genome towards comprehensive high-resolution analysis of genomic diversity and structural complexity. npj Genomic Medicine 2026.

[25] Warner JP, Barron LH, Goudie D, et al. A general method for the detection of large CAG repeat expansions by fluorescent PCR. 1996; 33:1022–1026.

[26] Schouten JP, McElgunn CJ, Waaijer R, Zwijnenburg D, Diepvens F, Pals G. Relative quantification of 40 nucleic acid sequences by multiplex ligation-dependent probe amplification. Nucleic acids research 2002; 30:e57.

