## Supplemental Table 1 for "Proximity-mapped whole-genome sequencing for characterization of repeat expansions and variants in highly homologous genomic regions"

Table S1. Conventional short-read WGS quality metrics for the 10 study samples.

| **Sample ID** | **Average autosomal coverage(X)** | **>20x Coverage (%)** | **Q30** | **Input Reads** | **Mapped Reads (%)** | **Unique Reads (%)** | **Proper Pairs (%)** | **Median  Insert Size** |
| --- | --- | --- | --- | --- | --- | --- | --- | --- |
| TP01 | 65.99 | 93.62% | 91.52% | 1,830 M | 96.53% | 91.01% | 92.71% | 216 |
| TP02 | 49.33 | 93.17% | 91.89% | 1,581 M | 95.67% | 84.51% | 91.44% | 194 |
| TP03 | 52.96 | 93.39% | 90.91% | 1,687 M | 96.91% | 84.36% | 93.19% | 188 |
| TP04 | 66.29 | 94.17% | 91.62% | 1,872 M | 96.41% | 90.83% | 92.28% | 208 |
| TP05 | 67.78 | 93.60% | 91.86% | 1,882 M | 96.43% | 91.70% | 92.84% | 201 |
| TP06 | 54.3 | 93.43% | 91.97% | 1,651 M | 96.14% | 87.77% | 92.20% | 189 |
| TP07 | 66.11 | 93.61% | 91.67% | 1,841 M | 96.75% | 91.80% | 93.08% | 200 |
| TP08 | 61.23 | 94.21% | 92.19% | 1,811 M | 96.26% | 86.99% | 91.75% | 217 |
| TP09 | 67.07 | 93.65% | 91.39% | 1,818 M | 96.81% | 92.54% | 93.20% | 212 |
| TP10 | 64.48 | 94.14% | 90.63% | 1,771 M | 96.65% | 92.91% | 93.02% | 201 |

Average autosomal coverage, mean sequencing depth across autosomes; >20x Coverage (%), percentage of bases covered at ≥20×; Q30, percentage of bases with Phred quality ≥30; Input Reads, total input sequencing reads; Mapped Reads (%), percentage of mapped reads; Unique Reads (%), percentage of uniquely mapped reads; Proper Pairs (%), percentage of properly paired reads; Median Insert Size, median fragment length of sequencing library.
