## Supplementary figures and images for "Proximity-mapped whole-genome sequencing for characterization of repeat expansions and variants in highly homologous genomic regions"

### Supplemental Figure 1

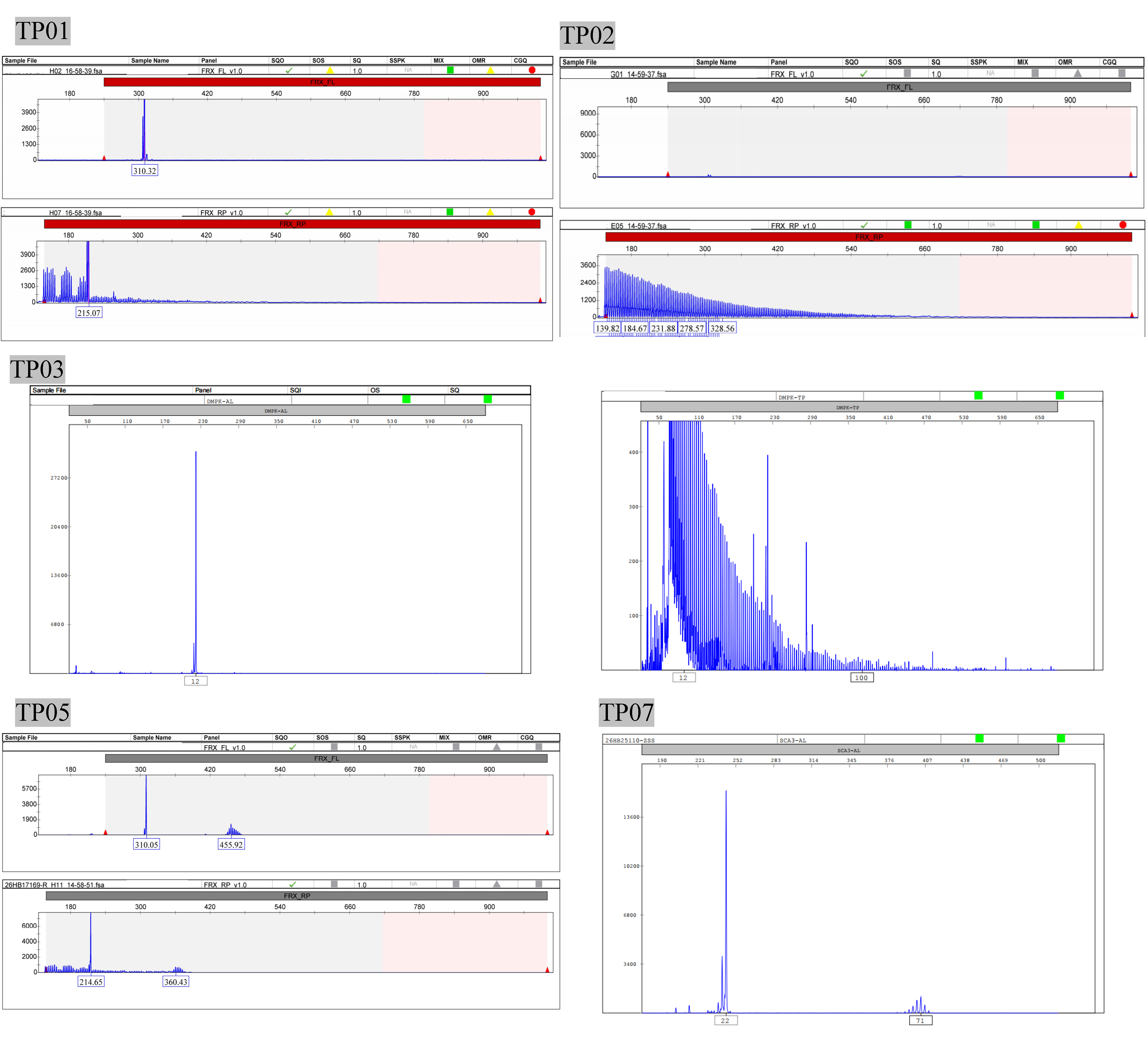

### Supplemental Figure 2

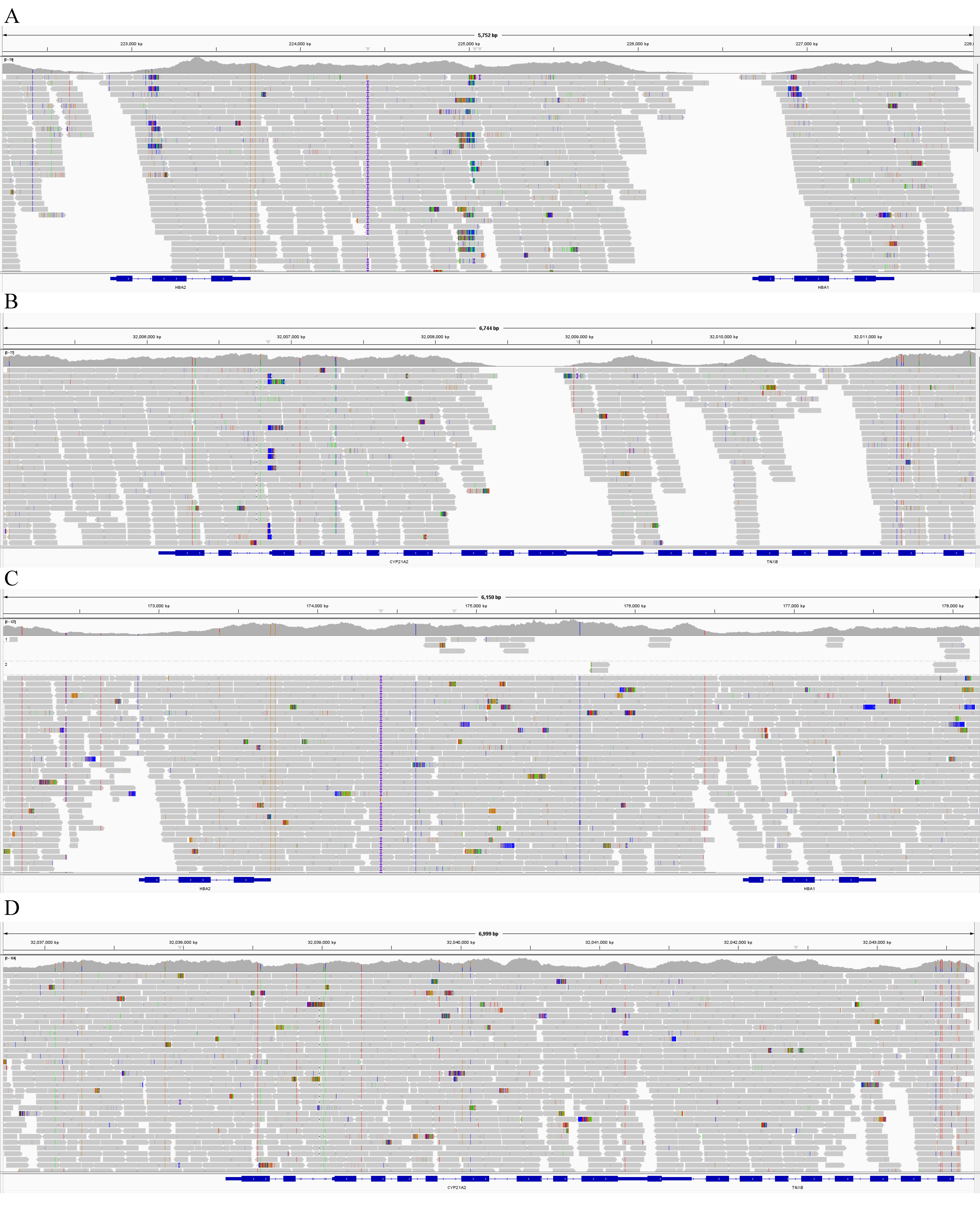
